# Prognostic Impact of Embryonal and Yolk Sac Components in Metastatic Germ Cell Tumors. Insights from an International Cohort

**DOI:** 10.64898/2026.02.10.26345982

**Authors:** Manuel Pedregal, Ignacio Mahillo, Isabel Miras, Begoña P. Valderrama, Rafael Morales-Barrera, David Humberto Marmolejo Castañeda, Nora Sobrevilla, Maria T. Bourlon, Praful Ravi, Victor Moreno, Christopher Sweeney

## Abstract

**Purpose:** Prognosis in metastatic non-seminomatous germ cell tumors (mNSGCT) is currently guided by the IGCCCG classification, which incorporates tumor markers, organs involved with metastatic disease, and primary site but not histologic subtype. We aimed to evaluate whether specific histological components provide additional prognostic information in a large international mNSGCT cohort.

**Patient and Methods:** We analyzed clinical, pathologic, and outcome data from 662 patients with mNSGCT across multiple international centers. Cox regression and multivariable stepwise models were used to evaluate the impact of age, tumor histology, serum markers, primary site of disease, chemotherapy, IGCCCG, and post-chemotherapy surgery on overall survival. Analyses were performed using both complete-case and imputed datasets to account for missing values.

**Results:** The presence of any percentage of embryonal carcinoma (EC) was independently associated with improved overall survival HR 0.603 (95% CI: 0.37–0.98, p=0.040), whereas yolk sac tumor (YST) predicted worse prognosis in complete-case analysis HR 2.27 (95% CI: 1.43 – 3.61 p = 0.001). Choriocarcinoma was also associated with a HR 1.58 (95% CI: 1.08 – 2.32 p= 0.019) adverse outcomes. IGCCCG risk classification remained a strong predictor of mortality HR up to 8.9 for Poor vs Good risk, (95% CI: 4.63 – 17.09 p < 0.001), but histologic components added significant independent prognostic value. Post-chemotherapy retroperitoneal lymph node dissection (RPLND) conferred a substantial survival benefit HR 0.44 (95% CI: 0.258 – 0.754 p=0.003). Interestingly, teratoma was not associated with mortality but was linked to younger age, testicular primaries, and higher likelihood of residual disease requiring surgery.

**Conclusions:** Histological composition, particularly the presence of EC or YST, has a significant and independent impact on survival in mNSGCT, beyond established risk classifications. Integration of histological subtypes may enhance prognostic accuracy and guide individualized treatment strategies in advanced germ cell tumors.

## INTRODUCTION

Testicular cancer represents the most common malignancy in young men and remains a model of curable metastatic disease owing to the extraordinary success of cisplatin-based chemotherapy.(1) The development of the International Germ Cell Cancer Collaborative Group (IGCCCG) classification provided a robust prognostic framework, stratifying metastatic non-seminomatous germ cell tumors (mNSGCTs) into good, intermediate, and poor categories with 5-year overall survival (OS) rates of approximately 90%, 80%, and 50–60%, respectively.(2,3) Despite this, substantial heterogeneity persists within each group, highlighting the limitations of a purely marker- and organ-based risk model and suggesting that additional biological features may influence outcome.

Histologically, NSGCTs comprise a range of elements derived from the pluripotency of embryonal carcinoma, with the potential to differentiate into yolk sac tumour (YST), choriocarcinoma, or somatic derivatives such as teratoma. These components vary in their chemosensitivity and natural history. (4) Teratoma is intrinsically resistant to systemic therapy, often necessitating surgical resection, yet its prognostic impact in the primary tumour remains disputed.(5,6) More recently, the IGCCCG Update Consortium provided the most comprehensive evidence to date, evaluating over 6,700 patients with mNSGCT, showing that the presence of teratoma (TER) in the primary tumour was associated with inferior PFS and OS in patients classified as good- and intermediate-risk, whereas no differences in outcomes were observed in those with poor prognosis.(7) This differential impact of teratoma depending on IGCCCG prognostic group may help to reconcile the conflicting results of two single-centre series. Memorial Sloan Kettering Cancer Centre, based on 193 patients, of whom 82 had teratoma in their primary tumour, reported an increased number of deaths in TER-positive patients; notably, only 40% received bleomycin, etoposide and cisplatin (BEP) and the analysis did not adjust for IGCCCG risk.(8) The more recent Indiana University study (1224 patients, 689 TER), similarly did not adjust for IGCCCG classification and had a median follow-up of 2.3 years, found no adverse prognostic impact of teratoma in orchiectomy specimens.(9)

In aggregate, these data suggest that histological composition may add prognostic value in mNSGCT, complementing established risk classifications and influencing surgical decision-making. We therefore sought to examine systematically the prognostic significance of histological subtypes in an international multicenter cohort treated predominantly in the modern BEP era, aiming to clarify the impact of histology on survival outcomes using uni- and multivariable Cox regression, with stepwise modelling applied to identify independent predictors beyond established prognostic factors.

## MATERIAL AND METHODS

### Patients

This retrospective, multicenter, international cohort study included male patients aged 18 years or older with histologically confirmed mNSGCT. Eligible patients were treated with cisplatin-based chemotherapy between 1992 and 2014 and had primary tumor specimens available for review. The specific details of the modern-dose platinum-based chemotherapy regimens administered to patients have been described in prior publications.(10–17) Patients were required to have sufficient clinical and pathological data for analysis. Exclusion criteria comprised incomplete data precluding outcome assessment, bilateral testicular cancer, primary retroperitoneal lymph node dissection (RPLND) before chemotherapy, or absence of retroperitoneal lymphadenopathy at baseline. Centers not able to provide the data at the requisite were excluded in their entirety.

Data was abstracted retrospectively from electronic health records at each participating center and entered into a secure centralized database. Baseline variables included age, IGCCCG prognostic group, serum tumor markers, primary tumor histology, primary site, and type and cycles of first-line chemotherapy. Treatment details included performance of post-chemotherapy RPLND or other surgical procedures. Pathological variables comprised the presence of viable carcinoma, YST, EC, choriocarcinoma, teratoma, and evidence of somatic malignancy. Histological components, including EC, YST, choriocarcinoma, teratoma, and seminoma—were documented when present in ≥1% of the overall tumor composition, as determined by local pathology review. Oncological outcomes included response to first-line therapy, relapse patterns, treatment at relapse, and survival status.

Follow-up data were obtained from clinical notes, radiological assessments, and death certificates where applicable. Vital status and date of last follow-up were recorded. Data quality was ensured through central consistency checks for completeness, missing values, and out-of-range entries; discrepancies were resolved with the responsible investigator. Site verification was not routinely planned but could be requested if systematic errors were identified. A flow-chart of patients included in this analysis is presented as **Figure 1**.

**Figure 1.**
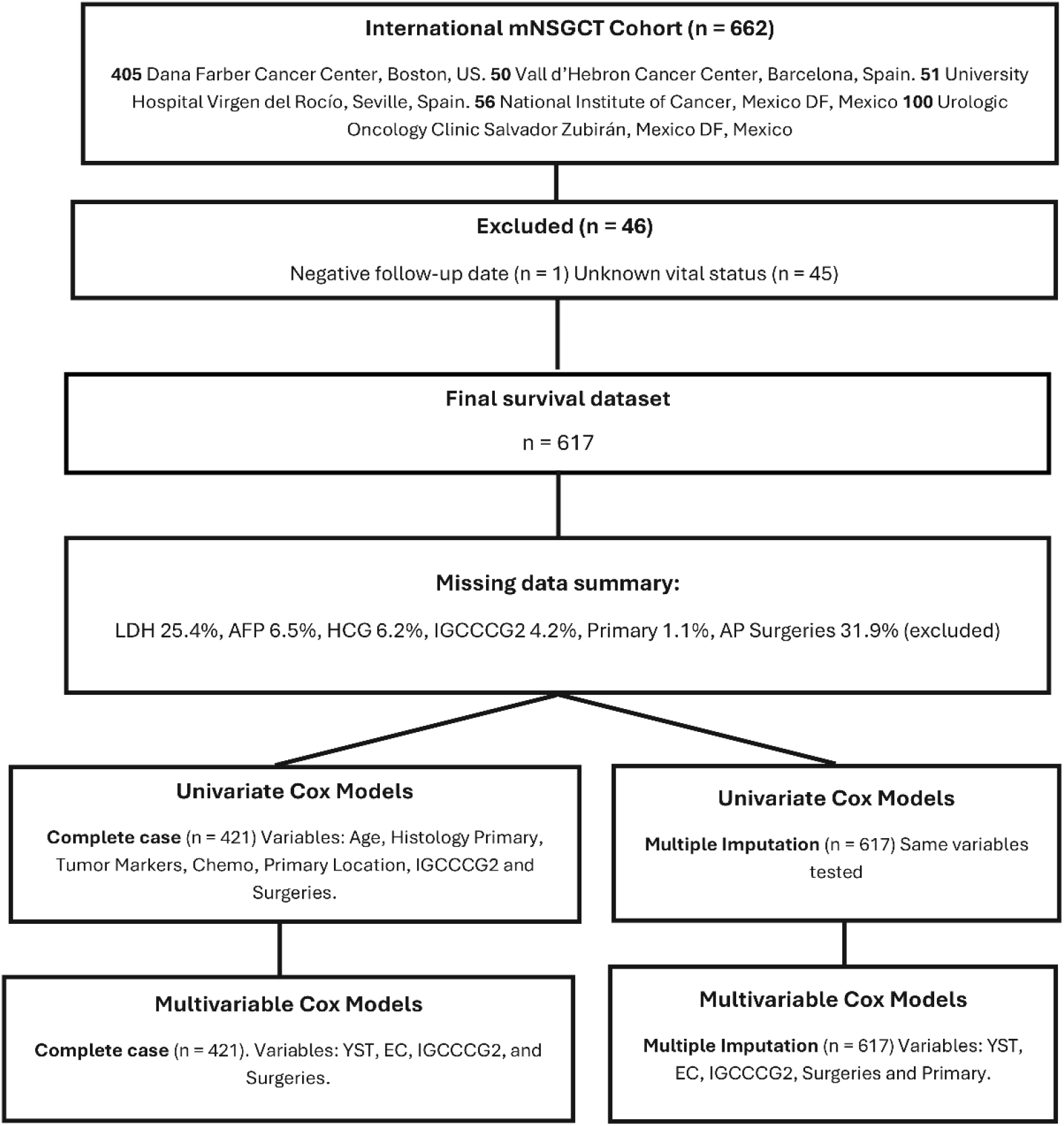
Consort diagram. Abbreviations: Chemo(chemotherapy), IGCCCG (International Germ Cell Cancer Collaborative Group), FU (follow up).

### Statistical Analysis

Patient’s characteristics were summarized using mean and standard deviation (SD) or median and interquartile range (IQR) for quantitative variables and counts and percentages for qualitative variables. Comparisons between the Teratoma and Non-Teratoma groups were performed using Student’s t test or Mann-Whitney U test for quantitative variables, and Chi-square or Fisher’s exact test for qualitative variables. Survival analysis was conducted to identify variables associated with mortality. Kaplan–Meier survival curves were estimated and compared using a two-sided log-rank test. Median survival times were calculated when applicable. Univariable and multivariable Cox regression models included the following variables: age; presence of any teratoma, yolk sac tumor (YST), seminoma, embryonal carcinoma (EC), or choriocarcinoma (CC); serum markers (AFP, HCG, and LDH); BEP chemotherapy; primary tumor location (testis, mediastinum, or retroperitoneum); IGCCCG risk group (good, intermediate, or poor); and post-therapy surgical approach (no surgery, RPLND, RPLND with additional procedures, or other surgery). Models were fitted using a stepwise procedure to derive the final multivariable model. Cox regression results were summarized using hazard ratios (HR), 95% confidence intervals (95%CI), and p-values (p). Additionally, Cox regression models were fitted using both the complete-case dataset and the dataset obtained through K-nearest neighbor imputation. A statistical analysis was performed using R version 4.2.1.

## RESULTS

### Patient Characteristics

A total of 662 patients with germ cell tumors were included in the cohort, with a median age of 29.9 years (SD ± 9.0). The primary tumor site was most commonly the testicle (90.0%), followed by the mediastinum (6.3%) and retroperitoneum (2.6%). At the time of analysis, 117 patients (17.7%) had died, while 500 (75.5%) were confirmed to be alive; vital status was unknown in 45 patients (6.8%).

Serum tumor markers at diagnosis were frequently elevated: alpha-fetoprotein (AFP) in 52.9%, human chorionic gonadotropin (HCG) in 52.9%, and lactate dehydrogenase (LDH) in 34.7% of patients. Risk classification according to IGCCCG was as follows: good-risk (52.1%), intermediate-risk (20.4%), and poor-risk (23.1%).

Histologically, YST, EC, choriocarcinoma, teratoma and seminoma components were identified in 272 (41.1%), 415 (62.7%), 180 (27.2%), 305 (46.1%) and 218 (32.9%) of cases, respectively. A total of 271 patients (40.9%) underwent retroperitoneal lymph node dissection (RPLND), and an additional 100 patients (15.1%) had RPLND in combination with other surgeries. The median follow-up time was 5.9 years (range, 0– 29.6). A detailed description of patient demographics and tumor characteristics is presented in **Table 1**.

**Table 1.** Patient and Disease Characteristics. Abbreviations: AFP, alpha-fetoprotein; BEP, bleomycin, etoposide, and cisplatin; CC, choriocarcinoma; EC, embryonal carcinoma; EP, etoposide and cisplatin; HCG, human chorionic gonadotropin; IGCCCG, International Germ Cell Cancer Collaborative Group; IGCCCG-2, updated IGCCCG risk classification; LDH, lactate dehydrogenase; NSGCT, nonseminomatous germ cell tumor; RPLND, retroperitoneal lymph node dissection; SD, standard deviation; TIP, paclitaxel, ifosfamide, and cisplatin; VeIP/VIP, vinblastine or etoposide, ifosfamide, and cisplatin; YST, yolk sac tumor. Values are presented as mean ± SD or number (percentage), as appropriate.

| **Variables** | **Value** |
| --- | --- |
| **Age** (mean ± SD) | 29.9 ± 9.0 |
| **Histology** |  |
| Teratoma – No | 357 (53.9%) |
| Teratoma – Yes | 305 (46.1%) |
| YST – No | 389 (58.8%) |
| YST – Yes | 272 (41.1%) |
| YST – Unknown | 1 (0.2%) |
| Seminoma – No | 441 (66.6%) |
| Seminoma – Yes | 218 (32.9%) |
| Seminoma – Unknown | 3 (0.5%) |
| EC – No | 246 (37.2%) |
| EC – Yes | 415 (62.7%) |
| EC – Unknown | 1 (0.2%) |
| CC – No | 482 (72.8%) |
| CC – Yes | 180 (27.2%) |
| **Baseline Vital Status** |  |
| Death – No | 500 (75.5%) |
| Death – Yes | 117 (17.7%) |
| Death – Unknown | 45 (6.8%) |
| **Primary Tumor Site** |  |
| Primary – Unknown | 7 (1.1%) |
| Primary – Testicle | 596 (90.0%) |
| Primary – Mediastinal | 42 (6.3%) |
| Primary – Retroperitoneum | 17 (2.6%) |
| **Risk classification (IGCCCG)** |  |
| IGCCCG – Good | 345 (52.1%) |
| IGCCCG – Intermediate | 135 (20.4%) |
| IGCCCG – Poor | 153 (23.1%) |
| **Tumor markers (AFP, HCG, LDH)** |  |
| AFP – Normal | 272 (41.1%) |
| AFP – Elevated | 350 (52.9%) |
| AFP – Unknown | 40 (6.0%) |
| HCG – Normal | 274 (41.4%) |
| HCG – Elevated | 350 (52.9%) |
| HCG – Unknown | 38 (5.7%) |
| LDH – Normal | 275 (41.5%) |
| LDH – Elevated | 230 (34.7%) |
| LDH – Unknown | 157 (23.7%) |
| **First-line chemotherapy regimen** |  |
| Chemotherapy – None | 8 (1.2%) |
| Chemotherapy – BEP-Based therapy | 569 (86%) |
| Chemotherapy – EP | 39 (5.9%) |
| Chemotherapy – VeIP/VIP | 34 (5.1%) |
| Chemotherapy – TIP | 4 (0.6%) |
| Chemotherapy – Others | 8 (1.3%) |
| **Surgical management (RPLND, others)** |  |
| Surgery – RPLND | 271 (40.9%) |
| Surgery – RPLND and others | 100 (15.1%) |
| Surgery – No surgery | 291 (44.0%) |
| **Residual pathology** |  |
| Pathology (residual) – Unknown | 237 (40.4%) |
| Pathology – Any teratoma | 150 (25.6%) |
| Pathology – Non-teratoma NSGCT | 58 (9.9%) |
| Pathology – Fibrosis | 133 (22.7%) |
| Pathology – Seminoma | 9 (1.5%) |

### Survival Analysis

Survival analysis was conducted in 617 patients after excluding those with unknown vital status or data inconsistencies. Kaplan-Meier curves demonstrated significantly worse overall survival in patients with YST histology (log-rank p < 0.019), mediastinal primaries (log-rank *p* < 0.001), elevated LDH (*p* < 0.001), or IGCCCG poor-risk classification (*p* < 0.001). Conversely, EC histology (log-rank p < 0.001) and RPLND (log-rank p < 0.002) were associated with improved survival. Estimated 5-year OS for patients who had embryonal component in the primary versus those who did not was 88.8% (95% CI, 81.1% to 90.4%) versus 73.4% (95% CI, 66.7% to 80.8%; P = .001; **Fig 2A)**, respectively. Regarding YST histology, the 5-year OS for patients who had YST in primary tumor versus those who did not was 78.7% (95% CI, 72.6% to 85.2%) versus 85.7% (95% CI, 81.1% to 90.4%; P = .019; **Fig 2B**). Analysis of mixed histologies showed that while embryonal carcinoma was associated with improved survival, the presence of yolk sac tumor conferred worse outcomes, with tumors lacking EC but containing YST showing the highest risk of death (EC−/YST+, HR, 3.62; 95% CI, 1.98– 6.61) and those containing both components demonstrating an intermediate prognosis (EC+/YST+, HR, 1.46; 95% CI, 0.74–2.86; log-rank P < .001), indicating that YST attenuates the survival advantage associated with EC in heterogeneous tumors.

**Figure 2.**
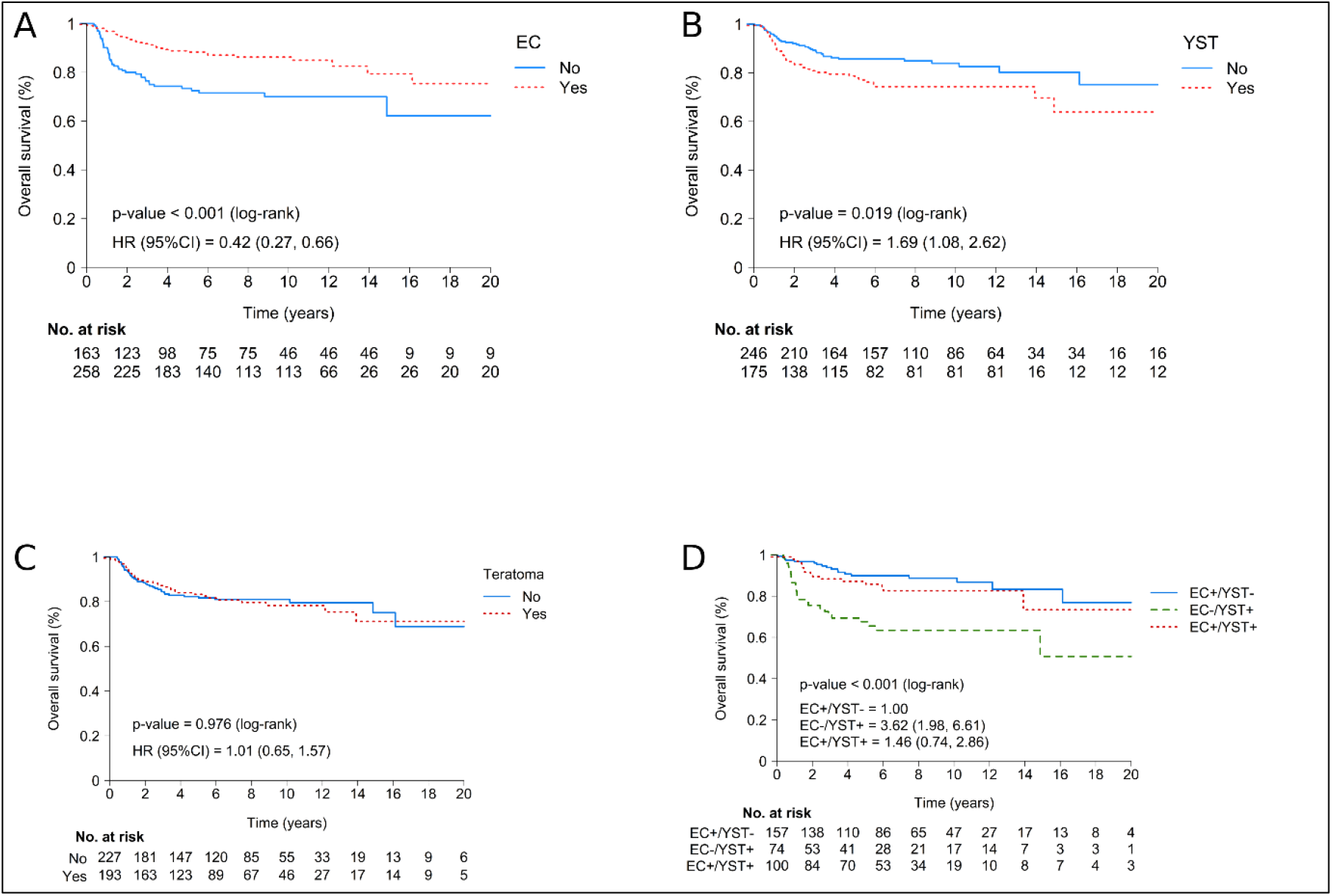
Kaplan–Meier overall survival curves according to histological components in metastatic non-seminomatous germ cell tumours. Panels show survival stratified by the presence of embryonal carcinoma (EC) (A), yolk sac tumour (YST) (B), teratoma (C), and mixed histology comprising both EC and YST components (D). Numbers at risk are shown below the x-axis where applicable. Abbreviations: CI, confidence interval; EC, embryonal carcinoma; HR, hazard ratio; OS, overall survival; YST, yolk sac tumor. P values were derived from two-sided log-rank tests. Hazard ratios and 95% confidence intervals were estimated using univariable Cox proportional hazards models.

Univariable Cox regression models were performed using both complete-case analysis (n = 421) and K-nearest neighbor imputation for missing values (n = 617) (**Table 2**). In both models, elevated LDH showed the strongest association with increased mortality (HR 3.07–3.54; *p* < 0.001). Elevated AFP (HR 1.68–1.86) and HCG (HR 1.72–1.75) were also independently associated with worse prognosis (*p* < 0.05 for all). EC histology was also associated with improved survival (HR 0.33–0.41; *p* < 0.001) in both models, while CC showed a negative impact in survival just in the imputed model HR 1.58 (95% CI: 1.08 – 2.32 p = 0.019). The IGCCCG intermediate- and poor-risk groups were associated with higher mortality compared to the good-risk group (HR 3.93–11.57; *p* < 0.001). Mediastinal primary site remained a significant adverse prognostic factor (HR ∼6.3–6.7; *p* < 0.001). RPLND was associated with improved survival (HR ∼0.43; *p* < 0.001), while other surgeries had no clear survival benefit. Kaplan-Meier curves stratified by IGCCCG risk, LDH level, and histology (YST, EC, seminoma) consistently mirrored the Cox regression results. Median overall survival was not reached for most subgroups given the relatively low event rate and long-term follow-up.

**Table 2.** Univariable Cox regression analysis. Abbreviations: HR, hazard ratio; CI, confidence interval; Ref, reference category. Hazard ratios were estimated using univariable Cox proportional hazards models. Two approaches are reported: a complete-case analysis including only patients with non-missing values, and a multiple-imputation analysis to account for missing covariates. For categorical predictors, the reference category is indicated as Ref (HR = 1.00). For continuous variables (e.g., age), HR describes the relative increase in mortality per one-unit increase. A p-value <0.05 was considered statistically significant.

|  | Complete-case analysis |  |  |  | Multiple imputation analysis |  |  |
| --- | --- | --- | --- | --- | --- | --- | --- |
|  | HR | (95%CI) | P |  | HR | (95%CI) | P |
| Age | 1.005 | (0.979-1.030) | 0.722 |  | 1.015 | (0.996-1.035) | 0.129 |
| Teratoma | 1.038 | (0.671-1.607) | 0.866 |  | 1.068 | (0.742-1.537) | 0.724 |
| YST | 1.715 | (1.107-2.656) | 0.016 |  | 1.202 | (0.834-1.733) | 0.324 |
| Seminoma | 0.603 | (0.360-1.011) | 0.055 |  | 0.669 | (0.442-1.013) | 0.058 |
| EC | 0.406 | (0.261-0.632) | <0.001 |  | 0.332 | (0.230-0.481) | <0.001 |
| CC | 1.411 | (0.896-2.224) | 0.137 |  | 1.582 | (1.079-2.319) | 0.019 |
| AFP | 1.857 | (1.152-2.993) | 0.011 |  | 1.677 | (1.135-2.478) | 0.009 |
| HCG | 1.725 | (1.070-2.780) | 0.025 |  | 1.747 | (1.179-2.587) | 0.005 |
| LDH | 3.069 | (1.941-4.853) | <0.001 |  | 3.535 | (2.406-5.194) | <0.001 |
| Chemo-BEP | 0.837 | (0.509-1.376) | 0.483 |  | 0.770 | (0.510-1.162) | 0.212 |
| Primary |  |  |  |  |  |  |  |
| Testicle | 1.000 |  |  |  |  |  |  |
| Mediastinal | 6.323 | (3.660-10.92) | <0.001 |  | 6.696 | (4.203-10.67) | <0.001 |
| Retroperitoneum | 2.022 | (0.733-5.577) | 0.174 |  | 1.843 | (0.676-5.029) | 0.232 |
| IGCCCG |  |  |  |  |  |  |  |
| Good | 1.000 |  |  |  |  |  |  |
| Intermediate | 3.930 | (1.954-7.905) | <0.001 |  | 4.629 | (2.581-8.303) | <0.001 |
| Poor | 9.430 | (5.099-17.44) | <0.001 |  | 11.57 | (6.951-19.24) | <0.001 |
| Surgeries |  |  |  |  |  |  |  |
| No surgery | 1.000 |  |  |  |  |  |  |
| RPLND | 0.441 | (0.258-0.754) | 0.003 |  | 0.434 | (0.272-0.693) | <0.001 |
| RPLND & others | 1.322 | (0.648-2.699) | 0.443 |  | 1.481 | (0.813-2.695) | 0.199 |

### Multivariable Analysis

Stepwise multivariable Cox regression identified IGCCCG risk classification, EC histology, YST, and RPLND as independent predictors of survival in both models **(Table 3)**. In the complete-case analysis, YST HR 2.27 (95%CI: 1.43 – 3.61 *p* = 0.001) and IGCCCG risk remained strong prognostic factors. EC histology HR 0.60 (95%CI 0.37 – 0.97 *p* = 0.040) and RPLND HR 0.29 (95%CI: 0.17 – 0.50 *p* < 0.001) were associated with improved survival. In the imputed dataset, age HR 1.03 per year (95% CI: 1.01 – 1.05 *p* = 0.007), EC HR 0.51 (95%CI: 0.34 – 0.77 *p* = 0.001), YST HR 1.52 (95%CI: 1.03 – 2.25 *p* = 0.034), and RPLND HR 0.31 (95%CI: 0.19 – 0.51 *p* < 0.001) retained independent prognostic significance. Mediastinal primary site also remained an adverse factor HR 2.02 (95%CI: 1.16 – 3.52 *p* = 0.013) in the imputed model. No significant interactions were observed between teratoma and IGCCCG risk, surgery, or histology in the multivariable framework.

**Table 3.** Multivariable Cox regression analysis. Abbreviations: HR, hazard ratio; CI, confidence interval; Ref., reference category. Hazard ratios were estimated using stepwise multivariable Cox proportional hazards models. Two models are presented: a complete-case analysis including only patients with non-missing covariates, and a multiple-imputation analysis accounting for missing data across predictors. For categorical variables, the reference category is indicated as Ref. For continuous variables (e.g., age), HR represents the relative change in mortality risk per one-unit increase. A p-value <0.05 was considered statistically significant.

| **Complete-case analysis** |  |  |  |
| --- | --- | --- | --- |
| Variable | HR | (95%CI) | P |
| YST | 2.272 | (1.428-3.615) | 0.001 |
| EC | 0.603 | (0.373-0.976) | 0.040 |
| IGCCCG |  |  |  |
| Good | Ref. |  |  |
| Intermediate | 4.134 | (2.017-8.473) | <0.001 |
| Poor | 8.897 | (4.632-17.09) | <0.001 |
| Surgeries |  |  |  |
| No surgery | Ref. |  |  |
| RPLND | 0.289 | (0.166-0.503) | <0.001 |
| RPLND and others | 0.781 | (0.369-1.652) | 0.518 |
| **Multiple imputation analysis** |  |  |  |
| Variable | HR | (95%CI) | P |
| Age | 1.026 | (1.007-1.046) | 0.007 |
| EC | 0.511 | (0.340-0.769) | 0.001 |
| YST | 1.522 | (1.032-2.244) | 0.034 |
| IGCCCG |  |  |  |
| Good | Ref. |  |  |
| Intermediate | 4.759 | (2.590-8.745) | <0.001 |
| Poor | 9.267 | (5.321-16.14) | <0.001 |
| Surgeries |  |  |  |
| No surgery | Ref. |  |  |
| RPLND | 0.310 | (0.189-0.507) | <0.001 |
| RPLND and others | 0.893 | (0.472-1.688) | 0.727 |
| Primary |  |  |  |
| Testicle | Ref. |  |  |
| Mediastinal | 2.021 | (1.160-3.521) | 0.013 |
| Retroperitoneum | 0.694 | (0.248-1.940) | 0.486 |

## DISCUSSION

We observed a favorable prognostic effect of embryonal carcinoma (EC), independent of IGCCCG risk classification. This result is biologically plausible: EC represents the least differentiated component of NSGCT, retaining pluripotency and an undifferentiated stem cell–like phenotype(4). It is characterized by intact apoptotic signaling, high mitochondrial priming, and pronounced chemosensitivity. Indeed, EC-dominant tumors frequently show marked responses to cisplatin-based chemotherapy, whereas differentiated components, particularly teratoma, are resistant. Our findings resonate with prior clinical observations and support the notion that EC, despite its aggressive clinical presentation, may serve as a marker of chemosensitive disease.

Conversely, yolk sac tumour (YST) was associated with inferior survival, even after adjustment for IGCCCG risk group. This aligns with earlier reports linking YST to late and chemoresistant relapses. Recent genomic studies provide mechanistic support: YSTs demonstrate distinct copy-number alterations, including recurrent amplifications of KRAS and KIT, deletions in ARID1A and PARK2, and overexpression of OVOL2, which has been implicated in cisplatin resistance. Importantly, YSTs retain wild-type TP53 but exhibit features of genomic instability, including microsatellite instability, which may further contribute to resistance(18). These molecular findings offer a plausible biological basis for the adverse clinical outcomes observed in our cohort, and emphasize the need for tailored therapeutic strategies, as conventional cisplatin-based chemotherapy may be insufficient for YST-predominant disease.

The role of teratoma in the primary tumor remains unsettled. In our series, it was not retained as an independent prognostic factor once IGCCCG and other covariates were included. This accords with the Indiana University cohort, which likewise found no effect, though interpretation was limited by a median follow-up of only 2.3 years and absence of IGCCCG stratification. In contrast, the Memorial Sloan Kettering report described an excess of disease-related deaths, but only 40% of patients received BEP, no adjustment for IGCCCG risk group was performed, and the relatively limited cohort size restricts the generalizability of the findings. In our cohort, 86% of patients received a BEP-containing regimen, and median follow-up was 5.9 years, providing a more mature dataset and treatment-homogeneous population for outcome assessment. While the substantially longer median follow-up of 17 years in the MSK series enhances sensitivity to very late relapse events, most late relapses occur within 4–5 years after treatment, a timeframe that is well captured in our cohort.

More recently, the IGCCCG Update reported that teratoma conferred inferior outcome in good-(5y-OS 94.5% vs 96.5%) and intermediate-risk patients (5y-PFS 76.9% vs 81.6%), while no difference was seen in the poor-risk group. Although they demonstrated an interaction between IGCCCG group and teratoma, the effect size was modest, with OS differences of only 2–3%. Given the substantially larger size of the IGCCCG Update Consortium cohort, it would be inappropriate to suggest that the present series refutes its conclusions regarding the association between teratoma and inferior outcomes in good- and intermediate-risk patients (7,19). Rather, our findings should be viewed as complementary. One plausible explanation for the modest adverse effect attributed to teratoma in the IGCCCG Update is that it may reflect, at least in part, the absence or underrepresentation of chemosensitive components, particularly embryonal carcinoma, rather than a strong independent biological effect of teratoma itself. In our analysis, the inclusion of embryonal carcinoma and yolk sac tumor as separate covariates revealed opposing prognostic signals, favorable for embryonal carcinoma and unfavorable for yolk sac tumor, which may attenuate the apparent contribution of teratoma when these components are not explicitly modeled. This observation suggests that the prognostic impact attributed to teratoma in large pooled analyses may be influenced by underlying histologic context heterogeneity and highlights the importance of considering individual histologic components jointly rather than in isolation. In our analysis, no such interaction was evident, suggesting that any adverse effect of teratoma is at most limited and might be overshadowed by established prognostic factors (20).

Our study also highlights the prognostic importance of surgical management. Patients undergoing post-chemotherapy retroperitoneal lymph node dissection (RPLND) experienced improved survival, although causality should be interpreted with caution. Several factors may contribute, including referral to high-volume centers with surgical expertise, lower burden of disseminated disease, and optimal clearance of residual teratoma or viable carcinoma (21). These findings align with longstanding evidence that RPLND performed in experienced centers reduces the risk of retroperitoneal relapses and improves long-term outcomes(22,23). Nevertheless, residual disease remains a challenge, particularly when surgery is incomplete or when somatic malignancy occurs within teratoma(24,25). Our results reaffirm the continuing importance of high-quality surgery as part of multidisciplinary management. Regarding the age, we did not replicate the adverse prognostic effect reported in the IGCCCG Update model, which likely reflects differences in sample size and cohort composition between studies rather than a true inconsistency with the IGCCCG findings.

This study has various limitations. To begin with, its retrospective and multicenter design introduces unavoidable heterogeneity in pathology assessment, surgical decision-making, and follow-up practices. In addition, although the cohort is large and contemporary, missingness in some variables required the use of imputation, which may not fully recapitulate true clinical patterns. The duration of follow-up was relatively short for a disease in which late relapses and late toxicities can occur. Finally, central pathology review was not performed, which in some cases may have led to disease misclassification, such as syncytiotrophoblast elements being incorrectly classified as choriocarcinoma (26). In addition, histological composition was based on local reports using a ≥1% cutoff, which may have resulted in misclassification of small tumor components, and metastatic histology data were unavailable, thereby limiting the evaluation of the impact of the histology on clinical outcomes and response to therapy.

Taken together, our findings reinforce the view that histological composition carries prognostic value in mNSGCT, beyond established IGCCCG stratification. While these findings are not intended to prompt immediate changes in standard clinical management, they may have practical implications. The adverse prognostic association of YST might support aggressive local control strategies, including surgery, analogous to current approaches in teratoma-containing disease, and more broadly, the distinct biological behavior of YST may guide drug development strategies to target YST biology related pathways (27). However, histology alone is unlikely to resolve the full granularity observed within prognostic groups. The integration of molecular with histological and clinical data could refine risk stratification and provide a path forward.

## Data Availability

All data produced in the present study are available upon reasonable request to the authors.

