## Supplementary figures and images for "Prognostic Impact of Embryonal and Yolk Sac Components in Metastatic Germ Cell Tumors. Insights from an International Cohort"

### Supplementary Material

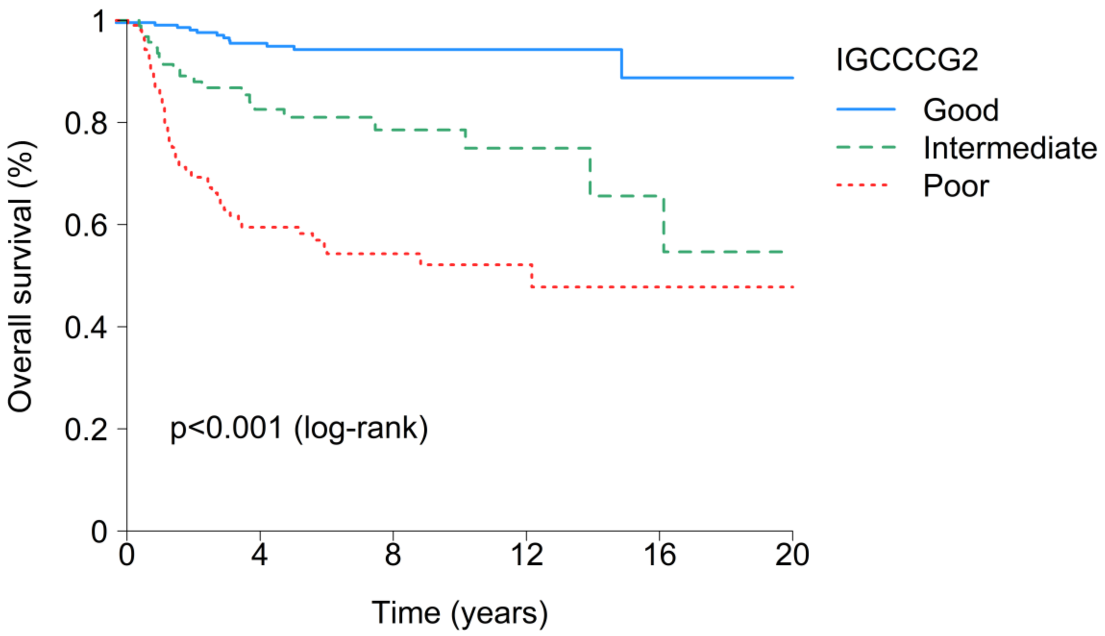

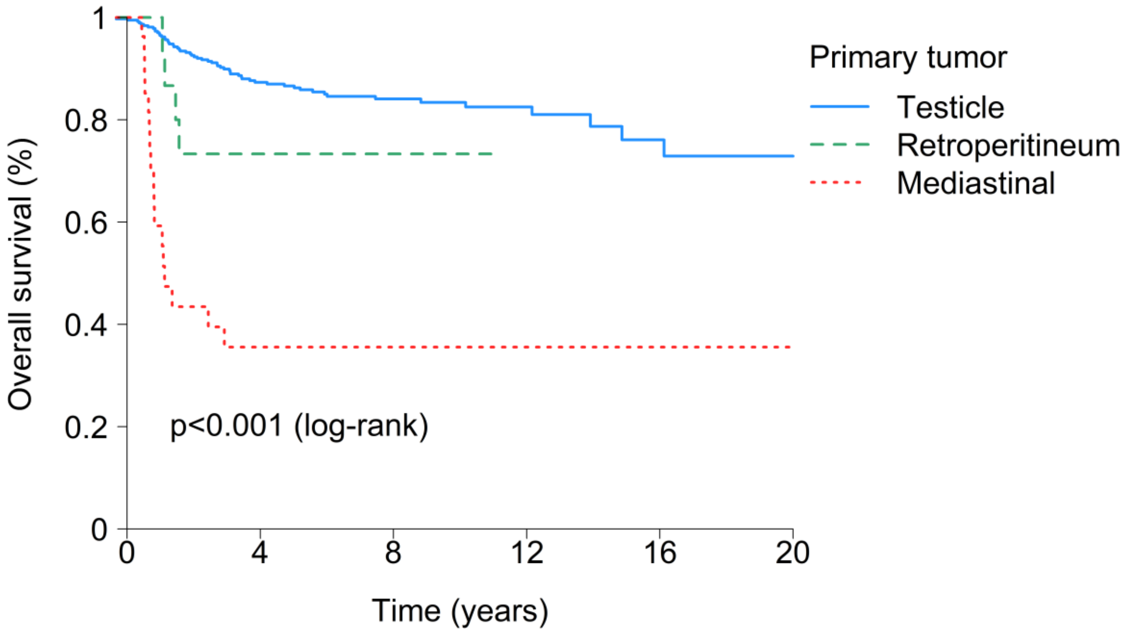

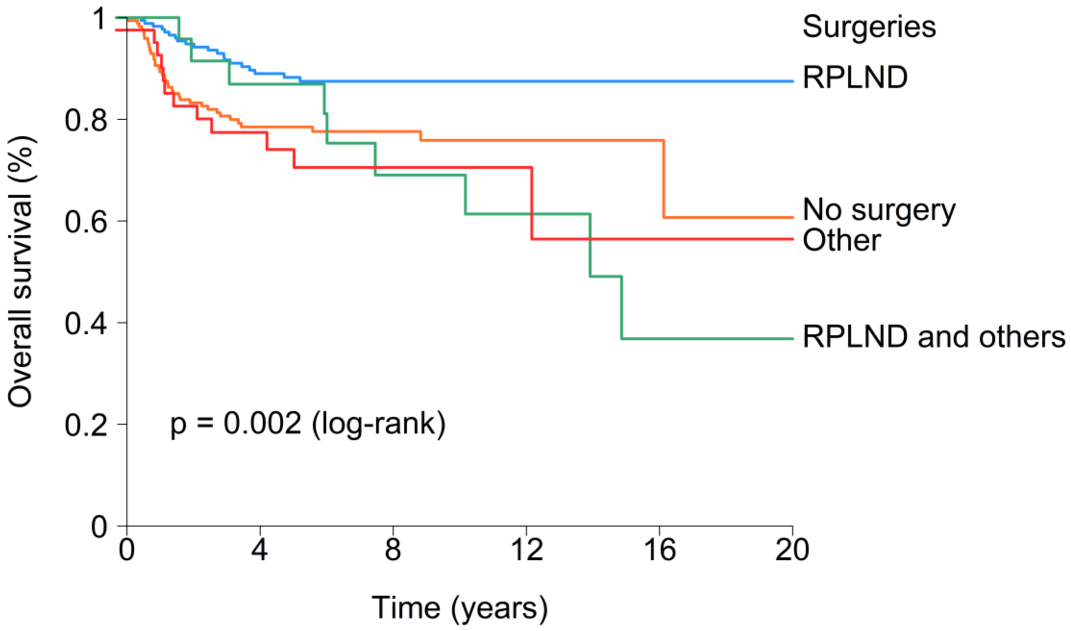

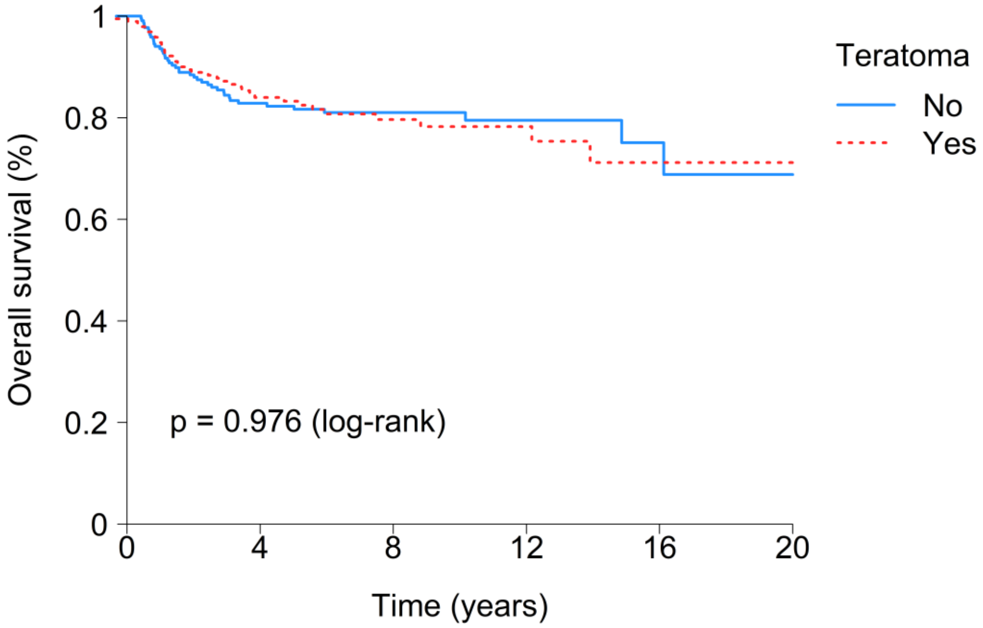

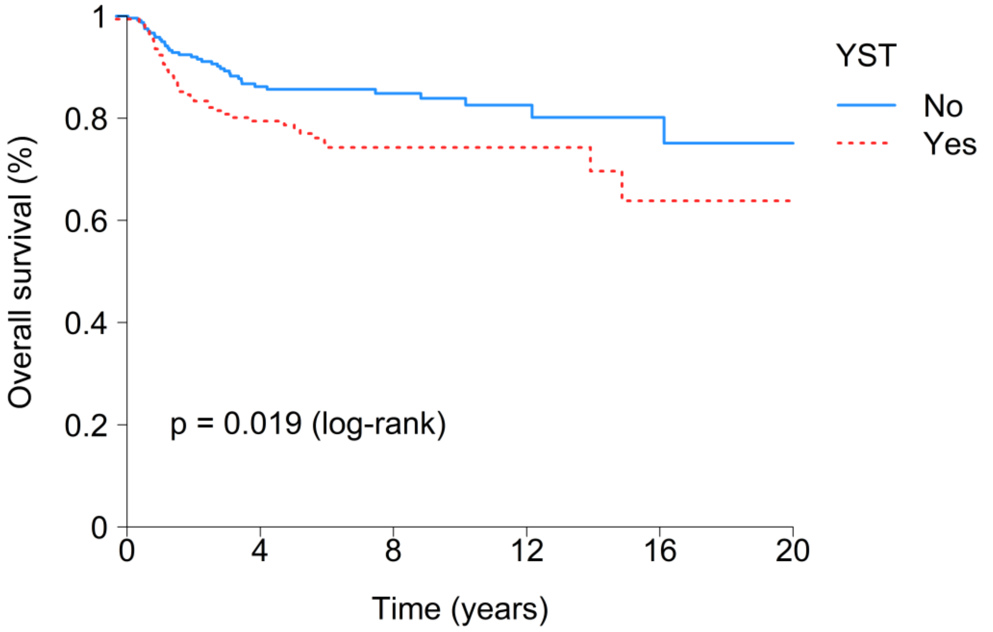

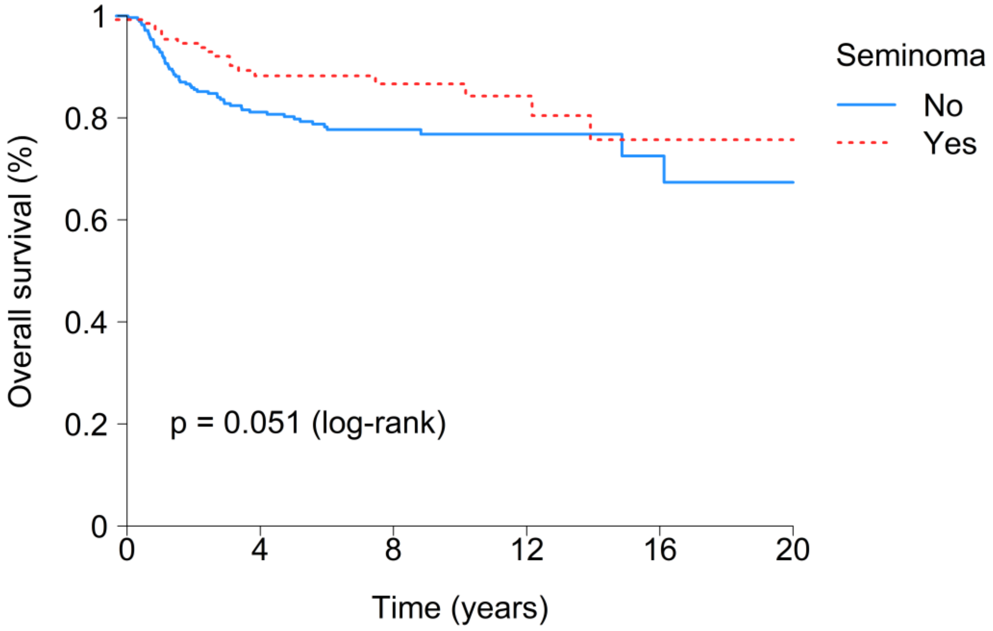

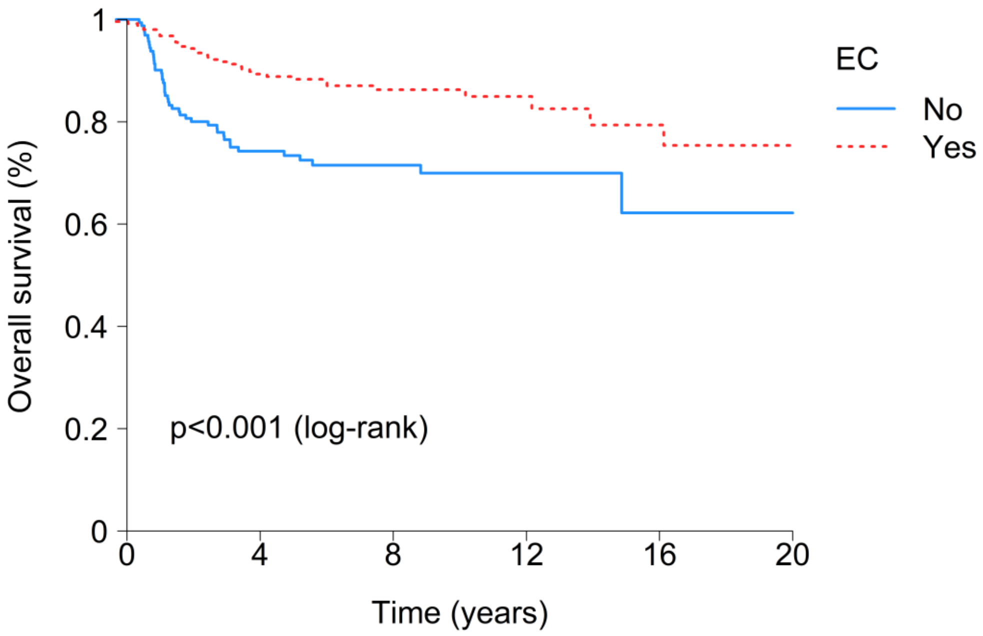

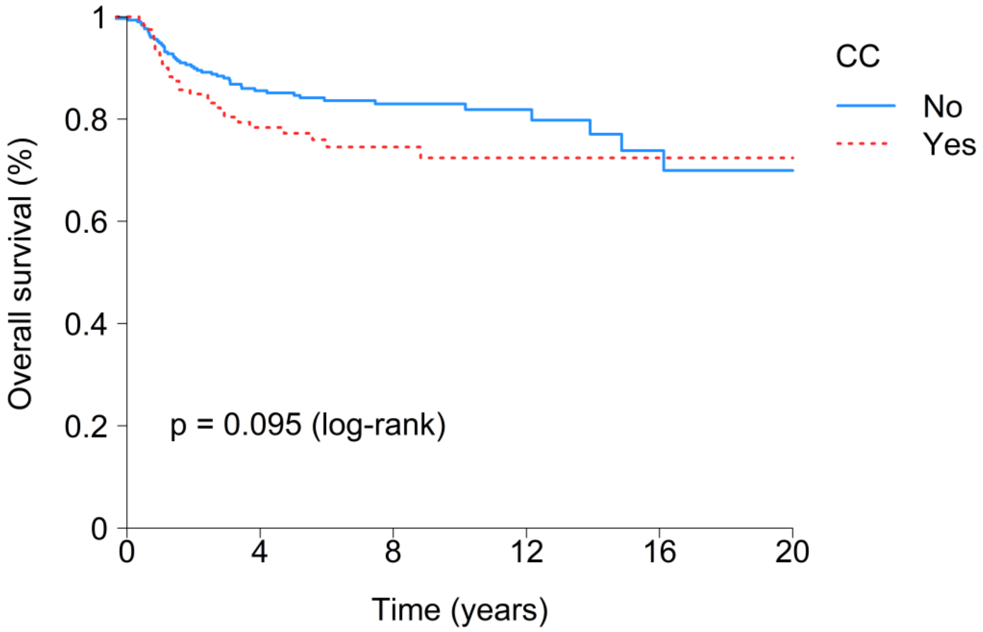

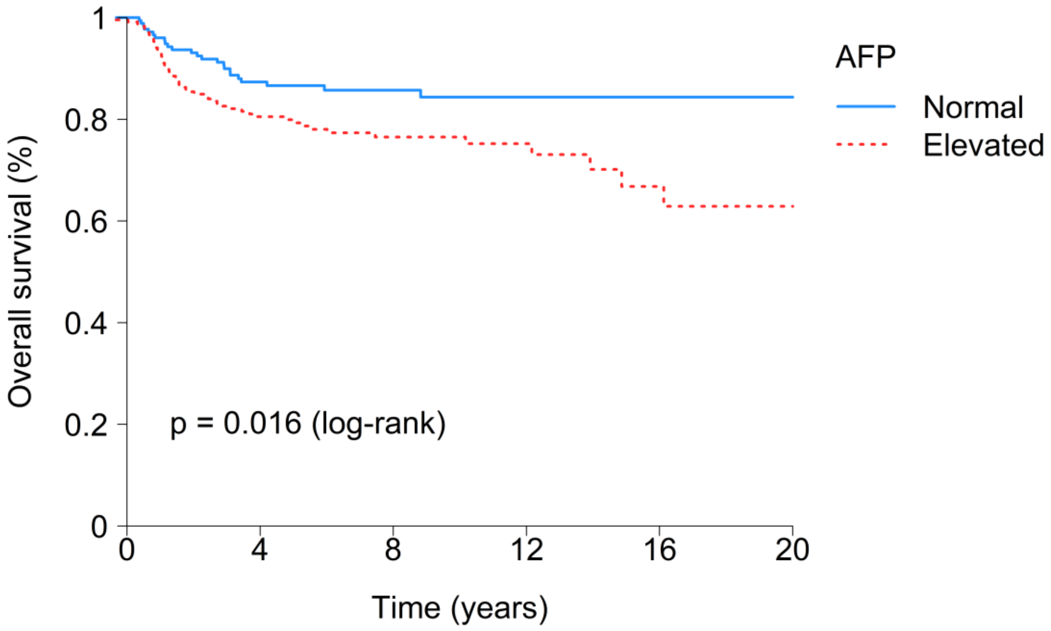

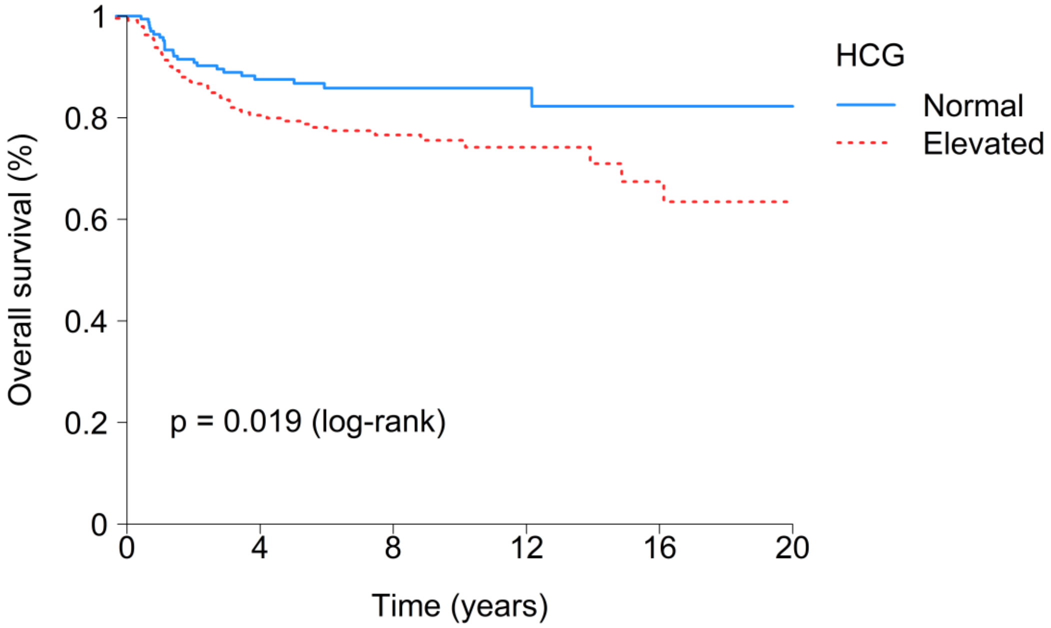

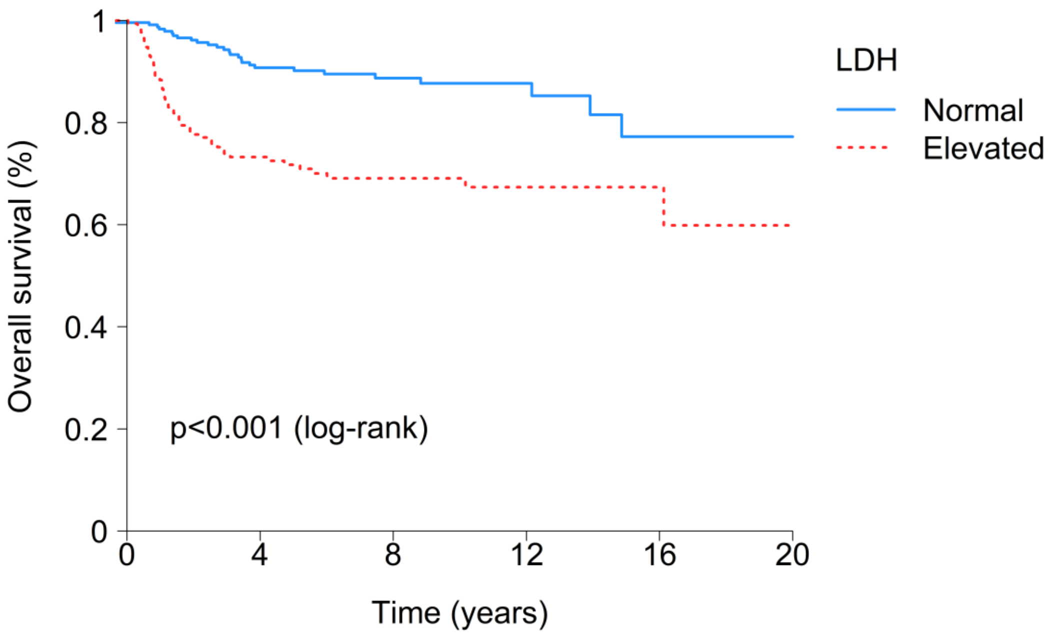

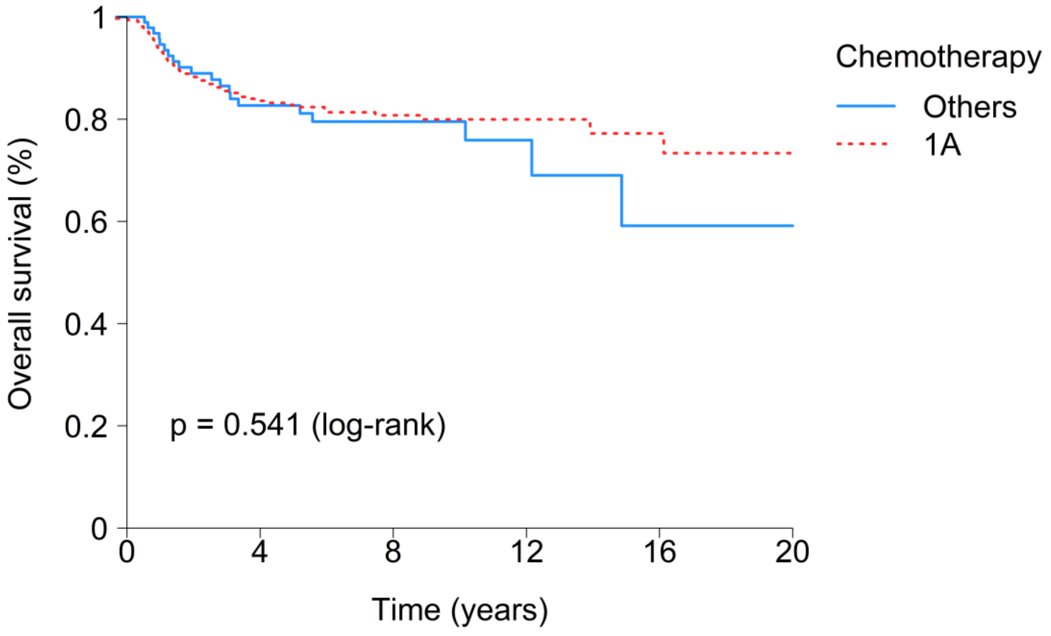

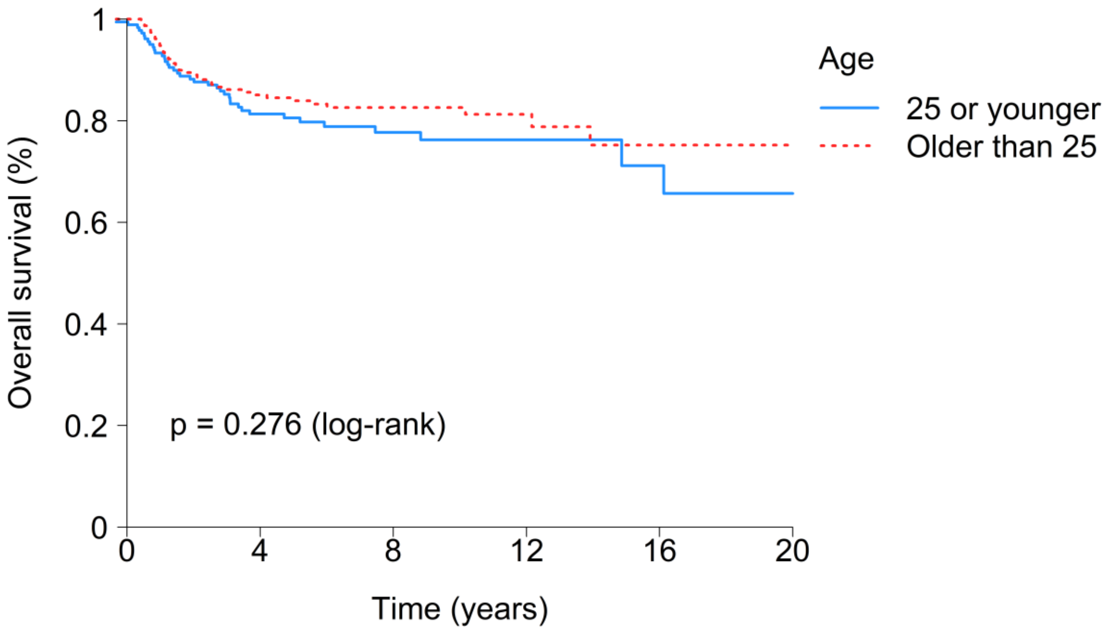
